# The Potential Impact of COVID-19 in Refugee Camps in Bangladesh and Beyond: a modeling study

**DOI:** 10.1101/2020.03.27.20045500

**Authors:** Shaun Truelove, Orit Abrahim, Chiara Altare, Stephen A. Lauer, Andrew S. Azman, Paul Spiegel

## Abstract

**Background:** COVID-19 could have even more dire consequences in refugees camps than in general populations. Bangladesh has confirmed COVID-19 cases and hosts almost 1 million Rohingya refugees from Myanmar with 600,000 concentrated in Kutupalong-Balukhali Expansion Site (age mean: 21 years, sd: 18 years, 52% female). Projections of the potential COVID-19 burden, epidemic speed, and healthcare needs in such settings are critical for preparedness planning.

**Methods and Findings:** To explore the potential impact of the introduction of SARS-CoV-2 in Kutupalong-Balukhali Expansion Site, we used a stochastic SEIR transmission model with parameters derived from emerging literature and age as the primary determinant of infection severity. We considered three scenarios with different assumptions about the transmission potential of SARS-CoV-2. From the simulated infections, we estimated hospitalizations, deaths, and healthcare needs expected, age-adjusted for the Kutupalong-Balukhali Expansion Site age distribution.

Our findings suggest that a large-scale outbreak is likely after a single introduction of the virus into the camp with 61-92% of simulations leading to at least 1,000 people infected across scenarios. On average, in the first 30 days of the outbreak, we expect 18 (95% prediction interval (PI), 2-65), 54 (95% PI, 3-223), and 370 (95% PI, 4-1,850) people infected in the low, moderate, and high transmission scenarios, respectively. These reach 421,500 (95% PI, 376,300-463,500), 546,800 (95% PI, 499,300-567,000) and 589,800 (95% PI, 578,800-595,600) people infected in 12 months, respectively. Hospitalization needs exceeded the existing hospitalization capacity of 340 beds after 55-136 days between the low and high transmission scenarios. We estimate 2,040 (95% PI, 1,660-2,500), 2,650 (95% PI, 2,030-3,380), and 2,880 (95% PI, 2,090-3,830) deaths in the low, moderate and high transmission scenarios, respectively.

Due to limited data at the time of analyses, we assumed that age was the primary determinant of infection severity and hospitalization. We expect that comorbidities and limited hospitalization and intensive care capacity may increase this risk, thus we may be underestimating the potential burden.

**Conclusions:** Our findings suggest that a COVID-19 epidemic in a refugee settlement may have profound consequences, requiring large increases in healthcare capacity and infrastructure that may exceed what is currently feasible in these settings. Detailed and realistic planning for the worst-case in Kutupalong-Balukhali and all refugee camps worldwide must begin now. Plans should consider novel and radical strategies to reduce infectious contacts and fill health worker gaps while recognizing that refugees may not have access to national health systems.

**AUTHORS’ SUMMARY:**

Why was this study done?
- Forcibly displaced populations, especially those who reside in settlements with high density, poor access to water and sanitation, and limited health services, are especially vulnerable to COVID-19.
- Bangladesh, which has confirmed COVID-19 cases, hosts almost 900,000 Rohingya refugees from Myanmar in the Cox’s Bazar district, approximately 600,000 of whom are concentrated in the Kutupalong-Balukhali Expansion Site.
- The capacity to meet the existing health needs of this population is limited; an outbreak of COVID-19 within this population threatens to severely disrupt an already fragile situation.
- We conducted this study to estimate the number of people infected, hospitalizations, and deaths that might occur in the Kutupalong-Balukhali Expansion Site to inform ongoing preparedness and response activities by the Bangladesh government, the United Nations agencies, and other national and international actors.

What did the researchers do and find?
- Using a dynamic model of SARS-CoV-2 transmission, we simulated how a COVID-19 outbreak could spread within the Expansion Site according to three possible transmission scenarios (high, moderate, and low).
- Our results suggest that a large-scale outbreak is very likely in this setting after a single infectious person enters the camp, with 0.5-91% of the population expected to be infected within the first three months and over 70-98% during the first year depending on the transmission scenario, should no effective interventions be put into place.
- Hospitalization needs may exceed the existing hospitalization capacity of 340 beds after 55-136 days of introduction.

What do these findings mean?
- A COVID-19 epidemic in a high population density refugee settlement may have profound consequences, requiring increases in healthcare capacity and infrastructure that exceed what is feasible in this setting.
- As many of the approaches used to prevent and respond to COVID-19 in the most affected areas so far will not be practical in humanitarian settings, novel and untested strategies to protect the most vulnerable population groups should be considered, as well as innovative solutions to fill health workforce gaps.

## INTRODUCTION

Severe acute respiratory syndrome coronavirus 2 (SARS-CoV-2) is responsible for hundreds of thousands of confirmed cases of COVID-19 globally to date. With growing concern about the global inability to control its spread, rapid preparation and planning are critical, particularly where this virus may have the greatest impact. More than 7.2 million people live in refugee camps and settlements worldwide, where high population density, limited water and sanitation infrastructure, and limited healthcare resources can create ideal conditions for the spread of infectious diseases.

Over 900,000 Rohingya refugees have sought refuge in Cox’s Bazar, Bangladesh; the majority arrived after 2017 fleeing ongoing violence in Myanmar (S1 Fig) [1]. Extending on flood-prone hilly terrain, the Kutupalong-Balukhali Expansion Site has 23 congested settlements with nearly 600,000 persons [2]. With over 46,000 persons per km^2^, this site could qualify as one of the densest cities on earth [3]. The majority of refugee families share their shelters, often composed by one single room. Half of the households have access to sufficient water to meet their needs [4]; all water and hygiene facilities are public and overcrowded [5]. The refugees are at high risk for epidemics given the high population density, poor health status, and limited health services. With confirmed COVID-19 cases in Bangladesh due to local transmission and ongoing transmission in neighboring countries, the introduction of the virus into the Expansion Site is likely.

The health status of refugees has improved since 2017 but remains fragile. The latest crude and child mortality rates are below emergency levels, while global acute malnutrition remains high [6]. The site is served by five hospitals run by non-governmental organizations (NGOs) and foreign governments with a total of 340 hospital beds (5.7 beds per 10,000 population) and up to 630 hospital beds when needed (10.6 per 10,000 population). There are 24 primary health care centers (1 PHC/25,000 persons) with numerous health posts, though the total number of functioning PHCs varies [7, 8]. Outside of the refugee site, there are 910 beds available in Cox’s Bazar district, including government, private, and NGO facilities for both the host community and refugees [9, 10]. The district hospital, with a 250-bed capacity, typically treats between 400-600 inpatients daily; 50-60 of whom are estimated to be refugees [10, 11]. It suffers from overcrowding with a bed occupancy rate over 200%, poor infection control, and inadequate hygiene protocol and waste management [10, 11]. While it has six intensive care unit (ICU) beds available, the ICU is reportedly not functional due to lack of trained staff and functional equipment [12]. However, the health cluster and its partners are planning to expand the health system capacity including increasing beds and staffing. Currently, there are an estimated 0.31 physicians and 0.12 nurses per 1,000 population in Bangladesh, far below the 4.5 skilled health workers per 1,000 population recommended by WHO [13, 14].

In this study, we aim to understand how SARS-CoV-2 might impact refugee camp populations, using the Rohingya refugees living in the Kutupalong-Balukhali Expansion Site as a case study. The primary aims of this analysis are to: 1) develop a baseline expectation of the possible infection burden, speed, and hospitalization capacity needed to respond to a COVID-19 epidemic; 2) use these findings to provide some recommendations to support ongoing preparedness planning by the Bangladesh government, United Nations agencies and other actors for a COVID-19 outbreak; and 3) apply lessons from this case study to refugees and other forcibly displaced persons globally.

## METHODS

We used a stochastic <u>S</u>usceptible <u>E</u>xposed <u>I</u>nfectious <u>R</u>ecovered (SEIR) mathematical model to simulate transmission in this population [15]. To capture the potential variability of transmission possible in this setting, we simulated epidemics under three potential scenarios with different values of the basic reproductive number, *R*_0_: 1) a low transmission scenario based on transmission levels in many of the Chinese provinces with elevated isolation and control practices and an *R*_0_ similar to influenza (*R*_0_=1.5-2.0) [16, 17]; 2) a moderate transmission scenario that mirrors estimates in early stages of the outbreak in Wuhan, China (*R*_0_=2.0-3.0) [18]; and, 3) a high transmission scenario where we assume that *R*_0_ is increased by a factor of 1.65 (*R*_0_=3.3-5.0) compared to estimates from other settings, as was observed during the 2017 diphtheria outbreak [19]. The *R*_0_ in each of these scenarios falls within the 95% confidence interval of the current range of estimates for COVID-19 [20]. We assumed an Erlang distributed serial interval (time between the onset of symptoms in infector-infectee pairs) with a mean of 6 days (standard deviation = 4.2) [21]. We assumed a population of 600,000 individuals and that the population was essentially closed (i.e., no movement, births, or deaths other than from COVID-19). Population characteristics and parameters used are detailed in Table 1, and further details about the model are in the Supplemental Information (**S1 Text**).

**Table 1.** Population characteristics and parameter values used in the model to estimate the transmission and impact of SARS-CoV-2 transmission in the Kutupalong-Balukhali Expansion Site.

| Parameter | Value (range) | Source |
| --- | --- | --- |
| **Population** |  |  |
| Size | 600,000 | UNHCR 2019 [2] |
| Median age | 16 years | UN WPP 2019[22] |
| Proportion female | *not accounted for* | - |
| **Transmission** |  |  |
| Basic reproductive number, $R_0$ | | |
| *Low transmission* | 1.5-2.0 | White et al. 2009[17] |
| *Moderate transmission* | 2.0-3.0 | Kucharski et al. 2020[18] |
| *High transmission* | 3.3-5.0 | Truelove et al. 2019[19] |
| Mean incubation period | 5.2 days | Lauer et al. 2020[23] |
| Infectious period | 6 days (standard deviation = 4.2) | Bi et al. 2020[21] |
| **Hospitalization and death** |  |  |
| Prop. of infections resulting in hospitalization | .036 (95% CI, .012-.093) | *estimated*<sup>1</sup> |
| Prop. of hospitalizations resulting in ICU admission | .32 | Huang et al. 2020[24] |
| Prop. of hospitalization resulting in death | .10 | Huang et al. 2020[24] |
| Time from onset to hospitalization | median: 3.42 days (sd: 0.79 days) | Bi et al. 2020[21] |
| Time from hospitalization to discharge | 11.5 days (95% CI, 8.0-17.3) | Sanche et al. 2020[16] |
| Time from hospitalization to ICU admission | median: 8.25 days (IQR, 4.8-14.0) | Zhang et al. 2020[25] |
| Time from hospitalization to death | 11.2 days (95% CI, 8.7-14.9) | Sanche et al. 2020[16] |
| Birth Rate | 0 | - |

<sup>1</sup>see **S1 Text** for details.

In addition to the population size and potential for elevated transmission (scenario 3), we also accounted for the age of this population in our estimates of severity and death, following consistent findings that age is a primary indication for severity and death. We estimated the probability that an infection results in severe disease with a logistic generalized addative model with a penalized cubic spline for age and a random effect for study, using data from five studies[21, 26-29]. We aggregated predictions from this model into estimates for 10-year age groups, and then applied those estimates to the age distribution of Kutupalong-Balukhali to get an overall probability that an infection that will develop into severe disease in this population (see **S1 Text** for more details). However, it should be noted our estimates of severe disease and death do not account for differences in comorbidities, differing attack rates by age due to population mixing characteristics, or various other factors.

We assume that in this setting hospitalization would be limited to those with severe disease (defined as tachypnea (≧30 breaths/ min) or oxygen saturation ≤93% at rest, or PaO2/FIO2 <300, and/or lung infiltrates >50% of the lung field within 24-48 hours) [30], and not used as a means of isolation. Thus, we assumed the proportion hospitalized was equivalent to the age-adjusted severe disease proportion calculated for the population and applied this to incident infections from the model simulations [21]. We assumed 32% of severe cases would require intensive care [24]. We estimated deaths assuming a 10% case fatality risk rate among hospitalizations/severe disease as was observed for SARS [31]. We conservatively did not account for potential increases in mortality when healthcare resources are exhausted. We assumed hospitalization occurs a median of 3.42 days after symptom onset (lognormally distributed, standard deviation=0.79), hospitalized cases are discharged after a mean of 11.5 days (95% confidence interval (CI), 8.0-17.3), and deaths occur after a mean of 11.2 days (95% CI, 8.7-14.9) [16].

## RESULTS

Our results suggest that a large-scale outbreak is likely in this population, even under the low transmission scenario, with 61% of the 1,000 simulations producing an outbreak of at least 1,000 people infected with a single introduction (Table 2) and increasing to 80% and 92% in the moderate and high transmission scenarios, respectively. On average, in the first 30 days of an outbreak following a single introduction, the estimated number of people infected ranges from 119 in the low transmission scenario to 504 in the high (Table 3). One year after the start of an outbreak, and in the absence of any effective interventions (e.g., vaccination, quarantine) or behavior change, between 73% (438,000/600,000; 95% prediction interval (PI), 62-81%) and 98% (591,000/600,000, 95% PI, 96-99%) of the population will have been infected (low and high transmission scenario) (Table 2).

**Table 2.**
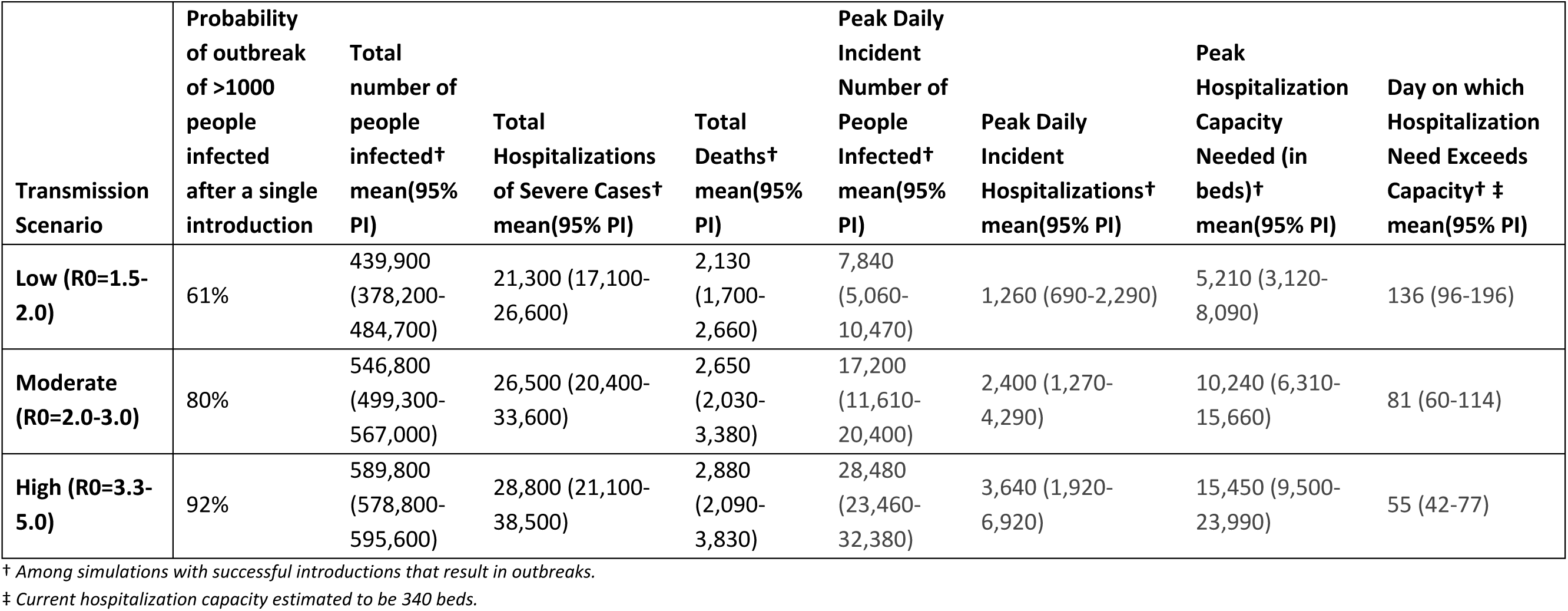
Overall impact of simulated outbreaks in Kutupalong-Balukhali Expansion Site, assuming no effective novel interventions or behavior change. *Total number of people infected, total hospitalizations*, and total *deaths* represent the final cumulative counts after the simulated outbreak has run its full course, among simulations that resulted in outbreaks of at least 1000 people infected. *Maximum daily incident* counts are the maximum incident counts of people infected and hospitalization on a single day across the full outbreak, occurring at the outbreak’s peak. We estimated the *daily hospitalization capacity* needed in beds using incident hospitalizations and assuming hospitalized individuals remain hospitalized for a mean of 11.5 days (95% CI, 8.0-17.3) [16]. We estimated the largest number of beds needed at any time over the course the outbreak, which we report as the *maximum capacity needed*. From the daily bed capacity needed over time, we also estimate the day on which this capacity needed exceeds the existing capacity of 340 beds, which we report as the *day on which hospitalization need exceeds capacity*.

| Transmission Scenario | Probability of outbreak of >1000 people infected after a single introduction | Total number of people infected† mean(95% PI) | Total Hospitalizations of Severe Cases† mean(95% PI) | Total Deaths† mean(95% PI) | Peak Daily Incident Number of People Infected† mean(95% PI) | Peak Daily Incident Hospitalizations† mean(95% PI) | Peak Hospitalization Capacity Needed (in beds)† mean(95% PI) | Day on which Hospitalization Need Exceeds Capacity† ‡ mean(95% PI) |
| --- | --- | --- | --- | --- | --- | --- | --- | --- |
| **Low (R0=1.5-2.0)** | 61% | 439,900<br>(378,200-484,700) | 21,300 (17,100-26,600) | 2,130<br>(1,700-2,660) | 7,840<br>(5,060-10,470) | 1,260 (690-2,290) | 5,210 (3,120-8,090) | 136 (96-196) |
| **Moderate (R0=2.0-3.0)** | 80% | 546,800<br>(499,300-567,000) | 26,500 (20,400-33,600) | 2,650<br>(2,030-3,380) | 17,200<br>(11,610-20,400) | 2,400 (1,270-4,290) | 10,240 (6,310-15,660) | 81 (60-114) |
| **High (R0=3.3-5.0)** | 92% | 589,800<br>(578,800-595,600) | 28,800 (21,100-38,500) | 2,880<br>(2,090-3,830) | 28,480<br>(23,460-32,380) | 3,640 (1,920-6,920) | 15,450 (9,500-23,990) | 55 (42-77) |

† Among simulations with successful introductions that result in outbreaks.
‡ Current hospitalization capacity estimated to be 340 beds.

**Table 3.** Cumulative number of people infected, hospitalizations, intensive care unit admissions, and deaths at 1, 3, and 12 months following successful introduction of simulations where an outbreak occurs.

| Transm-<br>ission<br>Scenario | 1 month |  |  |  | 3 months |  |  |  | 12 months |  |  |  |
| --- | --- | --- | --- | --- | --- | --- | --- | --- | --- | --- | --- | --- |
|  | Number of<br>People<br>Infected | Hospital-<br>izations | Intensive<br>Care Unit | Deaths | Number of<br>People<br>Infected | Hospital-<br>izations | Intensive Care<br>Unit | Deaths | Number of<br>People<br>Infected | Hospital-<br>izations | Intensive Care<br>Unit | Deaths |
| **Low**<br>($R_0=1.5-2.0$) | 18 (2-65) | 1 (0-3) | 0 (0-1) | 0 (0-0) | 2,127 (37-10,655) | 71 (1-374) | 6 (0-30) | 3 (0-15) | 421,500<br>(376,300-463,500) | 20,500<br>(16,700-25,000) | 6,500 (5,400-8,000) | 2,040 (1,660-2,500) |
| **Moderate**<br>($R_0=2.0-3.0$) | 54 (3-223) | 1 (0-6) | 0 (0-1) | 0 (0-0) | 125,838 (1,920-411,000) | 4,000 (48-16,700) | 183 (1-990) | 123 (1-631) | 546,800<br>(499,300-567,000) | 26,500<br>(20,400-33,600) | 8,500 (6,500-10,800) | 2,650 (2,030-3,380) |
| **High**<br>($R_0=3.3-5.0$) | 370 (4-1,850) | 6 (0-28) | 0 (0-1) | 0 (0-1) | 542,800<br>(260,300-594,300) | 25,000<br>(5,900-37,200) | 4,440 (98-10,100) | 1,880 (94-3,440) | 589,800<br>(578,800-595,600) | 28,800<br>(21,100-38,500) | 9,200 (6,700-12,300) | 2,880 (2,090-3,830) |

In all scenarios, we observed relatively slow growth during the beginning of the simulated outbreaks (Fig 1), with limited numbers of people infected and few, if any, hospitalizations and deaths during the first month after the introduction of one infectious case (Table 3). However, this quickly changed once sufficient infections were in the population, with cases rapidly increasing and the outbreaks culminating within the year (Table 3, Fig 1). By the time the first hospitalization occurs, we expect 50 (95% PI, 1-197) individuals to be infected in the population under the low scenario, assuming homogeneous probability of infection by age. This increases to 72 (95% PI, 2-289) and 141 (95% PI, 3-502) in the moderate and high scenarios, respectively. When the first hospitalization occurs, we expect the virus to have been circulating in this population for an average of 38, 30, and 23 days under the low, moderate, and high transmission scenarios, respectively.

**Fig 1.**
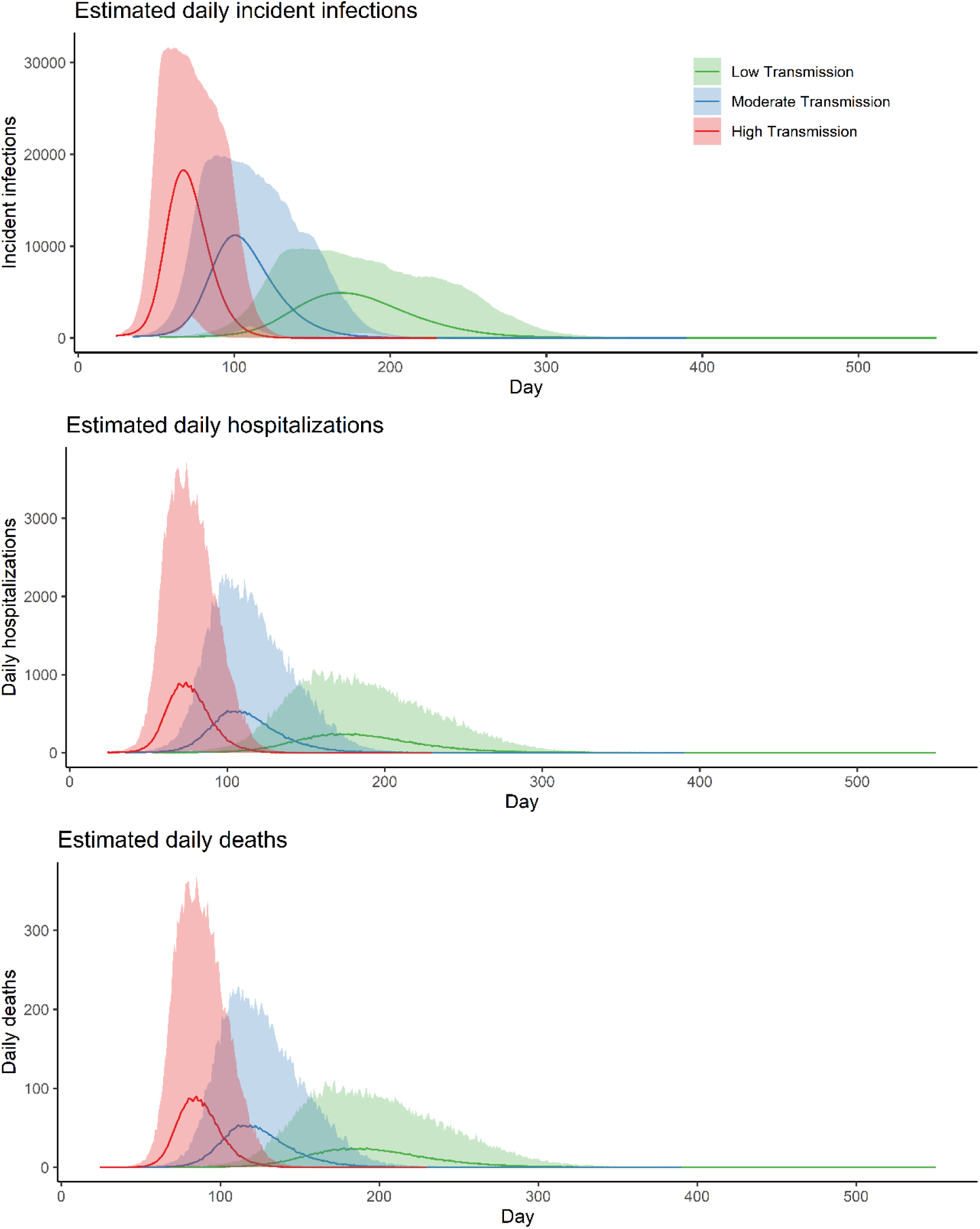
Simulated outbreak trajectories for Kutupalong-Balukhali Expansion Site camps under three transmission scenarios: low transmission (R0 =1.5-2.0), moderate transmission (R0=2.0-3.0), and high transmission (R0=3.3-5.0). (A) daily incident number of people infected by COVID-19, (B) daily incident hospitalizations, and (C) daily deaths under the three scenarios. The solid lines represent the mean outbreak trajectories and the shading represents the 95% prediction intervals of each scenario.

Adjusted for the age distribution in the Kutupalong-Balukhali Expansion Site, we estimated that 4.8% (95% CI, 0.3-15%) of infections in this population would result in severe disease and hospitalization. The maximum daily hospitalization capacity needed ranges between 5,200 (95% PI, 3,120-8,090) in the low transmission scenario and 15,450 (95% PI, 9,500-23,990) beds in the high (Table 2). Under the low transmission scenario, hospitalization needs exceeded the hospitalization capacity of 340 beds after 136 days (95% PI, 96-196 days) while in the high transmission scenario, this occurred after only 55 days (95% PI, 42-77 days; Table 2, Fig 2). Within 3 months of successful introduction, we estimated 6 (95% PI, 0-30) cumulative ICU admissions in the low transmission scenario compared to 4,400 (95% PI, 98-10,100) in the high transmission scenario (Table 3).

**Fig 2.**
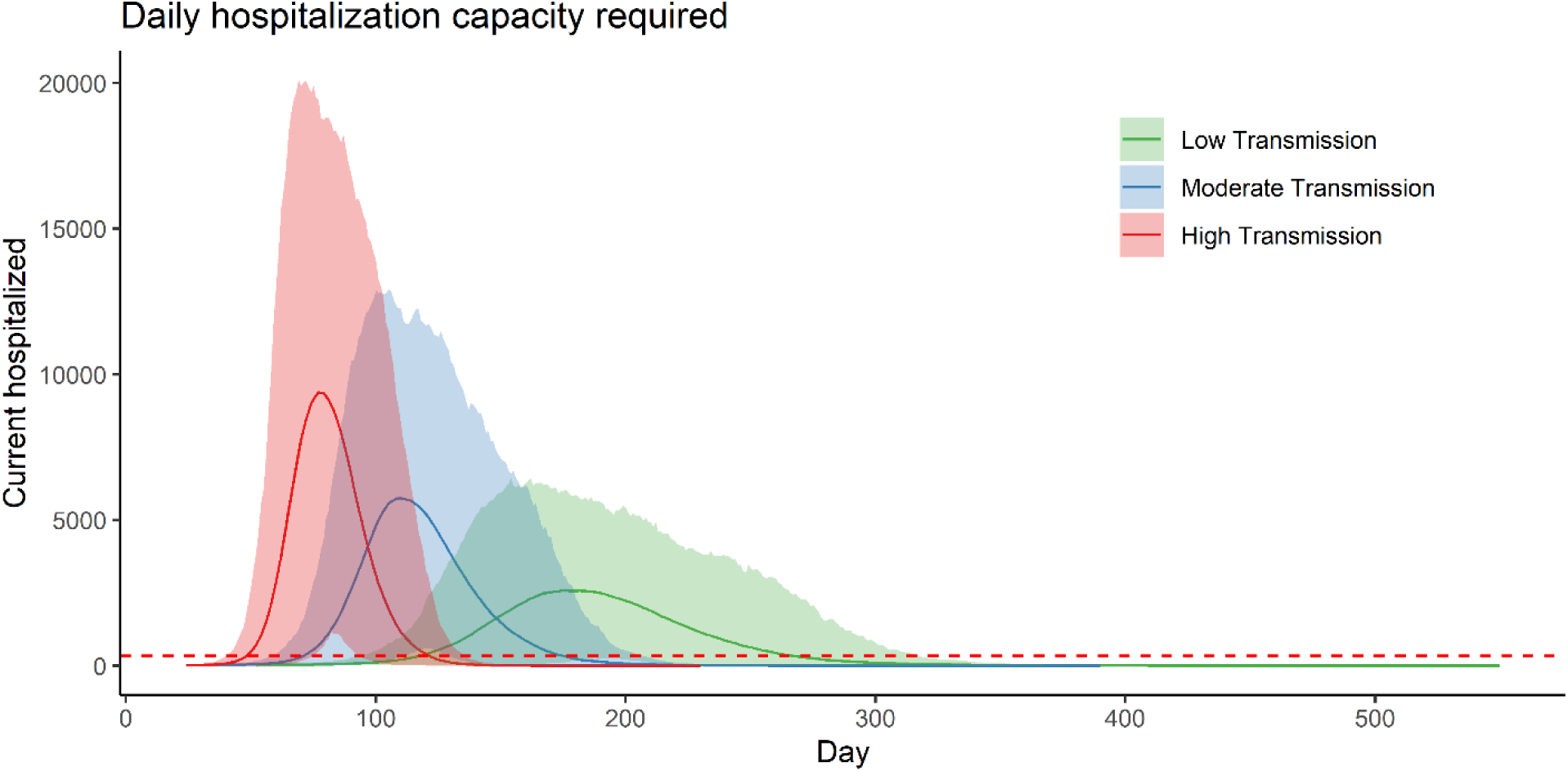
Hospitalization capacity requirements for an outbreak of SARS-CoV-2 in the Kutupalong-Balukhali camps, under three transmission scenarios: low transmission (*R*_0_ =1.5-2.0), moderate transmission (*R*_0_=2.0-3.0), and high transmission (*R*_0_=3.3-5.0). The solid lines represent the mean outbreak trajectories and the shading represents the 95% prediction intervals of each scenario. The dashed red line represents the 340-bed surge capacity currently believed to exist in the population.

Assuming that 10% of severe cases result in death, which puts the infection fatality rate at 0.48% (95% PI, 0.03-1.5%), we estimated between 2,130 (95% PI, 1,700-2,660) and 2,880 (95% PI, 2,090-3,830) deaths from the low to high transmission scenarios (Table 2). In the high transmission scenario, the maximum number of daily hospitalizations is reached on day 80, compared to day 117 and day 190 on average in the low and moderate scenarios, respectively (Fig 1).

## DISCUSSION

The Kutupalong-Balukhali Expansion Site has 23 congested settlements with nearly 600,000 persons residing there. Despite numerous challenges to the health of the population, including diphtheria and measles outbreaks, the health response and the resilience of the Rohingya refugees has been strong. However, the situation remains precarious and a COVID-19 outbreak in these camps would overload an already fragile health system and poor baseline health status. Using a stochastic disease transmission model, we estimated the number of people infected, hospitalizations, and deaths across three transmission scenarios after a successful introduction of SARS-CoV-2 into the Kutupalong-Balukhali Expansion Site. The introduction of SARS-CoV-2 into the Kutupalong-Balukhali Expansion Site or any other large refugee or internally displaced persons (IDPs) camp or settlement is likely to have serious consequences and overwhelm existing health systems. Even when transmission rates were assumed to be similar to that of influenza (low scenario), the necessary hospitalization capacities far exceeded the available capacities for the refugees in the Expansion Site in most simulations. Recent experience with other infectious disease outbreaks shows that the transmission of SARS-CoV-2 will likely be more intense in contexts such as Kutupalong-Balukhali Expansion Site than in out-of-camp settings because of high population density, inadequate water and soap to maintain hygiene, limited ability to isolate infected individuals, and large household sizes [19].

The younger demographic profile of the Rohingya camp population could reduce the overall rate of severe cases as compared with China or other upper-middle and high-income countries that have older populations. After adjusting for age, we estimate the overall severe disease rate among infections in this population (median=5.1%, [95% CI, 0.6-15.7%]) to be approximately half that estimated for China (median=10.1% [95% CI, 1.8-31.7%]). However, it is likely that co-morbidities such as malnutrition, concomitant diseases, and poor overall health status could cause more severe outcomes among these groups, which was not captured by our model.

Existing inpatient facilities in the camps are already operating close to full bed occupancy, and all scenarios show that they will be overwhelmed. Under current conditions, we expect this to occur within 2 to 4.5 months (55-136 days) of the first introduction of the virus, with cumulative hospitalizations reaching 71 to 25,000 in the first 3 months, depending upon the scenario; these numbers rise dramatically by 12 months. Current hospital bed surge capacity (630) as reported from the site may almost double the number of available beds, but even then, it only delays overwhelming the capacity by 3-10 days on average (S2 Table). Furthermore, while we have focused on the health needs of severe cases, health centers will be flooded with patients with mild to moderate symptoms also seeking care.

While we were unable to find an accurate recent estimate of the human resources currently available in the camps (e.g., number of doctors, nurses, midwives), it is extremely likely that they will be inadequate to face a large influx of patients. Given the limited availability of skilled health workers and hospital beds in Cox’s Bazar, it is unlikely that an additional influx of health care professionals within Bangladesh would be available. A surge of international health care workers could be considered, but this will be difficult given the high demands worldwide during the pandemic and existing travel restrictions.

Furthermore, issues of introduction of the virus from expatriates into Bangladesh would need to be carefully addressed. A more likely strategy will be task shifting among existing health care workers, with physicians treating the most severe cases, nurses the less severe cases, and community health workers addressing mild infections. Such task shifting would require intensive training that needs to begin immediately. An examination of the "re-purposing" of people working in various capacities (e.g., teachers as schools are closed) needs to be undertaken to assess how they can be used to address this epidemic while ensuring integration and complementarity of the response. Prepositioning of PPE and comprehensive training that is constantly reinforced to reduce health care worker exposures is critical to avoid sickness and death among this essential cadre. In particular, the community health workers will have to adapt their method of working to reduce chances of getting infected or infecting others. This may require using texts and calling people in the Rohingya dialect to ensure culturally appropriate communication and education. Furthermore, community health workers can help build trust among the Rohingya that contact tracing and isolation are necessary, and that they can have confidence in these services, given the long history of mistrust among the Rohingya and authorities in Rakhine state, Myanmar [32].

Given the limited number of beds for treating the predicted large number of severe COVID-19 cases in the Expansion Site, isolation through hospitalization, as is being done in other settings, will likely be very difficult. Alternative plans for the isolation of mild symptomatic infections are needed and are currently being discussed with the government of Bangladesh to help control such an outbreak, as was done during the 2017 diphtheria outbreak. Cholera treatment centers and diphtheria outbreak centers that are currently on standby are a low hanging fruit for repurposing. Setting up inclusive and accessible temporary hospitals to triage mild cases in these populations will require significant support and coordination by the government, UN agencies, and NGOs and may be limited by physical availability of land. Therefore, detailed advanced planning of healthcare capacities, triage procedures, and isolation strategies need to be finalized and shared widely as soon as possible.

Novel and previously untried strategies for social distancing and quarantine need to be considered. While culturally difficult and requiring strong socialization, isolating people over 60 years of age and those designated medically vulnerable together, so-called shielding, could be considered. One possibility could be isolating such groups among a group of families and relatives who have confidence in one another [33]. Consistent monitoring of fever and other symptoms combined with appropriate testing will be an integral part of this strategy. Any such shielding must be decided upon wholly by the community after much discussion. It would require sufficient resources to provide comprehensive care (e.g., food and supportive services) for the isolated community members. Since physical distancing will be extremely difficult, the use of facemasks among those most at risk, and possibly the whole population could be considered if there are eventually sufficient supplies. Rapid and creative solutions for improving hand hygiene, such as the installation of a multitude of handwashing stations and distribution of hand sanitizer combined with communication campaigns, will be needed. Access to accurate and consistent health information will be critical, as already rumors and misinformation have spread among Rohingya refugees about COVID-19, partly due to camp-wide restrictions on the internet and telecommunications [4]. In the future, the use of people who have recovered from COVID-19 infections will need to be considered after more data become available as to their ability to become reinfected and transmit to others, as was done with Ebola survivors [34]. The COVID-19 specific programs suggested above will require a strong surveillance system with adequate testing, case finding and isolation, as well as other supplies such as PPE. Such systems and supplies are not yet feasible in the camps, but will be essential to address the epidemic in this setting.

The three scenarios show varying degrees of increased mortality. Mortality due to COVID-19 is dependent upon various factors, particularly access to hospitals with ICUs and ventilators. Currently, there is only one facility with few mechanical ventilators in the camps and one facility with less than 10 ICU beds and no ventilators. Therefore, it is likely that mortality rates due to COVID-19 will be significantly higher here than in settings where nationals or refugees have such access. Reportedly, the ICUs and ventilators are not functioning in the district hospital in Cox’s Bazar, and thus access to health care facilities to treat severe cases of COVID-19 may be equally challenging for nationals. Mortality in Kutupalong-Balukhali Expansion Site is currently comparable to estimates for Chittagong division and nationwide, and are below emergency levels. However, the scenarios show a significant increase in mortality, ranging from 2,040 deaths (3.4/1,000/year) to 2,880 deaths (4.8/1,000/year) at 12 months due to COVID-19 alone; this does not take into account expected deaths that would have occurred during this time as well as increased deaths due to other illnesses that are untreated due to the outbreak.

As in other major epidemics where healthcare capacity and access to it is already limited, major outbreaks like this can easily disrupt an already precarious health system [35]. Diversion of these limited health resources from existing health services, including vaccination, obstetrical care, and emergency care, may cause an increase in mortality due to disease that could normally be treated by the health system; this occurred in the Ebola outbreak in West Africa where more people died from malaria than Ebola, and in Eastern DRC, where more people died from measles than Ebola [36, 37]. Such an increase in non-COVID-19 mortality is particularly concerning with the upcoming monsoon season in Bangladesh.

There are several limitations to this study. We are using a mass-action model, which tends to overestimate the size of outbreaks since populations are generally not closed and well mixed. However, due to population density and often closed nature of refugee camps, transmission in these settings has been shown to act more closely to theory. Additionally, the current evidence on the natural history and key epidemiological properties of COVID-19 reflect the interactions of the SARS-Cov-2 virus with non-displaced populations, and even in those populations many of these parameters remain poorly defined. Population structure and population health can lead to widely different burden of disease with no modifications to the virus. With the particular demographic characteristics and health status of refugee populations, like this one in Cox’s Bazaar, we need to be cautious when developing guidance based on previously estimated properties of SARS-CoV-2/COVID-19. Clinical surveillance, laboratory confirmation, and documentation are key to generating new evidence specific to this population and potentially generalizable to other refugee settings. Thus, we used the existing data that were available from documents and personal communications.

While we are focused here on the possible impact of SARS-CoV-2 on the Kutupalong-Balukhali Expansion Site, most of these findings are applicable to other refugee and IDP camp-like situations. While governments’ preparedness and response plans for COVID-19 may mention forcibly displaced populations, it is the details or lack of, that must be examined. The Inter-Agency Standing Committee recently released interim guidance for scaling up the COVID-19 response in camps and camp-like settings that highlights inclusion, protection, and readiness; and has since developed preliminary multi-sectoral preparedness and response plans [38]. This guidance, which applies the WHO guidance on COVID-19 preparedness and response [39] to populations in humanitarian crises, should be acknowledged in government and humanitarian agency decision-making.

Clear and detailed COVID-19 plans that explicitly state how existing programs will be adapted according to their context need to be developed and shared immediately. For example, different methods of food and fuel distribution will need to be undertaken that address social distancing issues. Potential consequences that isolation and other adaptive programming may cause, such as an increase in intimate partner violence, physical and sexual abuse among unaccompanied minors and other vulnerable groups, and mental health issues must be planned for in a concrete manner. Such adaptive programming will be an iterative process, as these are exceptional circumstances, and we will all have to learn by trying different approaches and then to document the results. A portal should be established by UNHCR and the global health cluster, led by WHO, to document how such adaptive programs for refugees and IDPs are being undertaken in different contexts. There will be a need for increased funding and basic supplies including PPE, as well as large-scale testing capabilities if these plans are to succeed.

The scenarios presented in this study focus on camp-like settings where refugee populations are relatively accessible, and health services are available and generally free. However, the majority of refugees reside outside of camps, often in urban contexts [40]. For these populations, availability, access, cost, and quality of health services vary by host government policies. These refugee populations are also particularly vulnerable to the serious consequences of pandemics as targeted responses among a dispersed group of refugees, and equity in health service delivery is very difficult to achieve.

During exceptional times, it is not unreasonable for governments to take extraordinary measures to protect their citizens; in fact, that is what is expected of governments. Therefore, while difficult for governments to state openly at this point, it is possible that many countries will restrict access to their hospitals to nationals only, particularly when there are large numbers of refugees in the same geographical area. We are already seeing governments closing their border to non-nationals, and in some cases, not allowing people to seek asylum [41]. Such a predicament leaves refugees and other non-nationals, such as undocumented migrants, in an extremely precarious position. During a pandemic, there will be limited support by countries to establish field hospitals with ICU capabilities for refugees in camp-like settings, as they will be occupied with addressing their own national response. Unfortunately, there is no simple recommendation as to how to address this life and death issue. However, we believe it is important to acknowledge the likelihood that host countries’ hospitals will be closed to refugees in most camp-like settings, and consequently, COVID-19 preparedness and response plans should proceed on realistic scenarios.

Finally, refugees face discrimination and are often falsely accused of spreading disease. The widespread rise of populism combined with anti-migrant and anti-refugee sentiments that we are observing globally provides a hostile environment that could be exacerbated by a pandemic. We are concerned that the COVID-19 pandemic, while completely unrelated to being a refugee, could be used as an excuse to take retribution against refugees, as well as other vulnerable groups such as IDPs and undocumented migrants. Such retribution could take many forms, including restricting or stopping asylum seekers, which is currently occurring, the closure of camps, and forced repatriation (refoulement, which is against refugee law), all used in the name of public health. Not only would this be morally wrong, it would jeopardize the effectiveness of containment and mitigation measures, as pandemics require planning and responses that do not discriminate by nationality and protect the health of the global population.

Our findings suggest a COVID-19 epidemic among the Rohingya refugees in the Kutupalong-Balukhali Expansion Site will be extremely difficult to mitigate. Many of the current interventions such as social distancing, contact tracing and isolation, good hygiene, and sophisticated treatment in ICUs for critical cases will be challenging to implement. How SARS-CoV-2 will transmit and how COVID-19 will manifest in such a population with a poor baseline heath status and acute malnutrition is simply unknown at present. Beyond preparing now for COVID-19 through specific and sensitive interventions for this population to reduce morbidity and mortality, there is a need to rigorously document what does occur when the epidemic affects such a refugee population so we can learn how to better prepare and respond in similar settings. This will require strong surveillance systems to be established now. It will require working closely with the community to better understand how social distancing can work for them in their situation. And it will require the continued commitment of the brave health care workers and the refugees themselves to respond to this pandemic.

## Data and Analysis Code

All code and data are provided on the repository https://github.com/HopkinsIDD/covidKutupalong.

## Data Availability

All code and data are provided on the repository https://github.com/HopkinsIDD/covidKutupalong.

## Supplemental Figures, Tables, and Text

**S1 Text**. Supplemental Information. Additional details on the methods of the model and the severity age-adjustment.

**S1 Table**. Comparison of selected indicators of health status and health service availability in Kutupalong-Balukhali extension site and Bangladesh.

**S2 Table**. Day on which hospitalization requirements exceed current estimated bed capacity (340 beds) and estimated surge capacity (630 beds) in the Kutupalong-Balukhali Expansion Site.

**S3 Table**. Model transitions and rate parameters used in the SEIR model.

**S1 Fig**. Bangladesh health facilities by type in Cox’s Bazar’s camp sites, Health Sector, Cox’s Bazar, March 2020. Source: UN Office for the Coordination of Humanitarian Affairs; https://www.humanitarianresponse.info/en/operations/bangladesh/health.

**S2 Fig**. (A) Age distribution comparison between Kutupalong-Balukhali Expansion Site, Bangladesh overall, and China, and (B) the estimated probability of severe disease given infection with SARS-CoV-2, by 10-year age group.

**S3 Fig**. Comparison of the expected overall proportion of infections that will result in severe disease, as estimated through adjustment for population age distribution.

